# Identifying Key Predictors of Smoking Cessation Success: Text-Based Feature Selection Using a Large Language Model

**DOI:** 10.1101/2025.06.18.25329854

**Authors:** Thuy T. T. Le, Jiongxuan Yang, Zimo Zhao, Kaidi Zhang, Wenjun Li, Yan Hu

**Affiliations:** University of Michigan School of Public Health, Department of Health Management and Policy, Ann Arbor, MI, USA; University of Michigan School of Public Health, Department of Biostatistics, Ann Arbor, MI, USA; The Chinese University of Hong Kong, School of Data Science, Shenzhen, China; University of Massachusetts Lowell, Department of Public Health and Center for Health Statistics, Lowell, MA, USA

## Abstract

**Background:** The most effective way to reduce mortality and morbidity among current smokers is to quit smoking. Although about half of smokers attempted to quit, only one-tenth succeeded in 2022.

**Objective:** To identify key predictors of smoking cessation success to inform cessation interventions and increase quitting rates.

**Methods:** We analyzed data from waves 5 and 6 of the Population Assessment of Tobacco and Health (PATH) study (December 2018 to November 2021). Using OpenAI’s GPT-4.1, we identified the top 45 variables from wave 5 that are highly predictive of 12-month smoking abstinence in wave 6, based on descriptions of survey variables. We then validated the predictive power of the GPT-4.1-selected variables by comparing the performance of eXtreme Gradient Boosting (XGBoost) trained on different sets of variables. Finally, we derived insights into the top 10 variables, ranked according to their SHapley Additive exPlanations values.

**Results:** The performance of XGBoost trained with all possible wave 5 variables and the 45 selected variables was almost identical (AUC:0.749 vs AUC:0.752). The top 10 variables included past 30-day smoking frequency, minutes from waking up to smoking first cigarette, important people’s views on tobacco use, prevalence of tobacco use among close associates, daily electronic nicotine product use, emotional dependence, and health harm concerns.

**Conclusion:** This study demonstrates the ability of OpenAI’s GPT-4.1 to identify the top 45 PATH wave 5 variables associated with 12-month smoking abstinence using only their descriptions. This approach could help researchers design more effective survey questionnaires and improve efficiency of data collection.

**What is already known on this topic:** Generative artificial intelligence models have recently been applied to assess their potential in addressing various tobacco-related issues, such as detecting tobacco products in social media videos and promoting vaping cessation. However, their application in identifying the most significant predictors of tobacco use behavior, based on survey data, remains unexplored.

**What this study adds:** GPT-4.1 successfully assigned high-quality importance scores to survey variables for predicting 12-month smoking abstinence over two years among current established smokers. It accomplished this using only the textual descriptions of the survey variables, without accessing the actual survey data. Based on these importance scores, GPT-4.1 can aid in identifying the most crucial variables for predicting smoking cessation success.

**How this study might affect research, practice or policy:** This study demonstrates the capacity of GPT-4.1 to perform feature selection, paving the way for future exploration of this innovative approach to address other tobacco-related issues.

## Introduction

Smoking cessation remains a public health priority due to the well-documented adverse health effects associated with tobacco use, including cardiovascular diseases, chronic obstructive pulmonary disease, and various forms of cancer [1, 2]. Despite substantial efforts, achieving sustained smoking cessation continues to be a significant public health challenge. In 2022, 67.7% of the 28.8 million US adults who smoked expressed a desire to quit, and 53.3% attempted to do so. However, only 8.8% successfully quit smoking [3]. This limited success highlights the complexity of the cessation process, driven by a multitude of biological, psychological, and social factors. Advanced research methodologies are needed to better identify the key predictors of cessation behaviors and improve the efficacy of future interventions.

Smoking cessation success is influenced by a complex interplay of various factors, including genetic [4, 5], psychological [6, 7], social and environmental elements [8-12], which interact in dynamic and multifaceted ways. Better understanding of the key predictors of cessation success is essential for enhancing quit rates. Traditional machine learning methods, such as eXtreme Gradient Boosting (XGBoost) and Random Forest, have demonstrated their efficiency in automatically identifying variables significantly associated with smoking behaviors within extensive datasets [13, 14]. These methods, however, require rigorous training on empirical data to be effective.

Unlike classical machine learning models, several recent studies have indicated the potential of large language models (LLMs) in performing feature selection based solely on textual descriptions of survey variables, without direct access to the dataset [15, 16]. The ability to pinpoint important variables predictive of tobacco use behaviors without conventional data access could be invaluable in the design of surveys. LLMs are deep learning models with a massive number of parameters that are pre-trained on vast amounts of data, allowing them to perform a variety of tasks such as language translation, text summarization, sentiment analysis, question answering, text generation, information retrieval etc. [17]. They have a wide range of potential applications across domains, including healthcare, medicine, engineering, biology, finance, marketing among others [17, 18]. By leveraging their advanced natural language processing capabilities, LLMs have the potential to transform data interpretation and inform decision-making processes in public health and clinical settings.

The Population Assessment of Tobacco and Health (PATH) study is a nationally representative longitudinal cohort study that collects comprehensive data on participant socio-demographic information, tobacco use patterns and habits, tobacco risk perceptions, and health status among others [19]. It is an invaluable resource for studying smoking cessation behavior. In this study, we investigate the use of a LLM – OpenAI’s GPT-4.1 - in identifying the most important predictors of smoking cessation success in the PATH data. We first employed GPT-4.1 to perform feature selection based solely on the text descriptions of the variables, without accessing the actual data. Then we used XGBoost to evaluate how well these GPT-4.1-selected features are at predicting smoking cessation success in 2 years using the PATH data. XGBoost is an ensemble learning method that builds multiple decision trees and integrates them to enhance prediction accuracy, while also handling large datasets efficiently [20]. By leveraging the robustness and efficiency of XGBoost, we seek to 1) assess the efficacy of GPT-4.1 for text-based feature selection, 2) gain deeper insights into the factors that facilitate or impede smoking cessation success. The pioneering use of LLMs to study smoking cessation behavior will pay the way for future exploration of this innovative approach to address other tobacco-related issues. This study’s findings could inform targeted interventions and policies designed to support individuals in their efforts to quit smoking, ultimately contributing to better health outcomes at the population level.

## Materials and Methods

### Data

For this study, we utilized data from the adult PATH survey [19] waves 5 (December 2018 to November 2019) and 6 (March 2021 to November 2021). The adult PATH study employs a four-stage stratified sampling design to collect data from individuals aged 18 and older, both tobacco users and non-users, who live in U.S. civilian, non-institutionalized settings [21]. The combination of these two waves provides us with a robust sample size for assessing smoking cessation success defined as 12-month smoking abstinence. The cohort included current smokers who had smoked 100 or more cigarettes in their lifetime and were smoking some day or every day at wave 5. Among 34,309 adults who completed wave 5, 25,743 (75%) also participated in wave 6. Of these, 6,120 (24%) were classified as current smokers in wave 5 and had a non-missing 12-month smoking status in wave 6. Smokers were considered to have successfully quit smoking if they had not smoked a cigarette in the 12 months leading up to the wave 6 survey. Among 6,120 smokers from wave 5, 496 (8%) successfully quit by wave 6.

We started with a merged wave 5 and wave 6 dataset that consists of 2,315 wave 5 variables, encompassing participants’ various social demographic characteristics, heath status, and tobacco-related information among others. We first filtered the data to include only current smokers from wave 5, removing not directly relevant variables such as survey weights, random variables, and those with more than 2.5% missing data or low variation. Missing values were imputed using mean for numerical variables and mode for factor variables. Subsequently, we eliminated highly correlated variables with a cutoff probability of 0.75 to further reduce the number of independent variables and enhance the performance of downstream models.

Our final dataset contains 6,120 wave 5 current smokers, of whom 496 individuals were abstinent from smoking for 12 months by wave 6, and 303 wave 5 variables, which were used for further analysis. Of these variables, we employed GPT-4.1, a state-of-the-art LLM from OpenAI [22], to select the top 45 most important variables from wave 5 in predicting smoking cessation success in wave 6. This selection was based solely on the textual descriptions of the variables, rather than their values. Subsequently, we trained XGBoost on the data with GPT-4.1-selected variables to evaluate the efficacy of GPT-4.1 in context of feature selection and gain insights into quitting behavior. The dataset was divided into training data (80%), which was used for fine-tuning XGBoost’s hyperparameters and training XGBoost, and test data (20%) which was exclusively for evaluating the performance of XGBoost on unseen data. We used GPT-4.1 to perform feature selection based solely on textual descriptions of survey variables, i.e., without access to the actual dataset.

### Feature Selection

To enhance the performance of XGBoost and decrease computational costs, we leveraged GPT-4.1 to conduct feature selection using prompting techniques to identity a subset of the most informative wave 5 variables that are highly predictive of smoking status in Wave 6. Specifically, we crafted prompts that presented the model with descriptions of PATH survey variables one at a time, using the model to assign a numerical score ranging from 1 to 100, with 1 being the least influential and 100 being the most influential, to reflect each variable’s importance in predicting smoking cessation status. Notably, while the model processed the descriptive text of each variable, it did not have direct access to any actual data within the dataset. Additionally, we did not provide the model with any survey details or context. We anticipated that the model’s extensive training would enable it to effectively rank these variables based on its pre-existing knowledge. Further details on our prompt design can be found in the Appendix. Due to random nature of LLMs, we requested GPT-4.1 to rank each variable 50 times and each variable’s the final score is the average of 50 scores to obtain robust rankings of these variables. The efficacy of GPT-4.1 in the context of text-based feature selection would be evaluated using XGBoost in the following section. For this task, we set the GPT-4.1 parameter *temperature* to 0.5 to balance coherence and creativity.

### Model Development

After GPT-4.1 provided a list of scores for 303 wave 5 variables, we trained XGBoost on the training data using (i) all 303 variables and (ii) the 45 highest-scored variables. To evaluate the performance of XGBoost on the test data in each case, we conducted a series of experiments with 1000 different splits of the training dataset into training and validation subsets. For each experiment, several hyperparameters of XGBoost [23] such as learning rate (*eta*), maximum tree depth (*max_depth*), minimum child weight (*min_child_weight*), subsampling (*subsample*), and column sampling (*colsample_bytree*) were optimized using Bayesian optimization [24] within predefined parameter ranges to enhance model performance, see Table A1 in the Appendix. These parameters helped balance learning speed, control model complexity, and ensure generalization by using subsets of features and data at each iteration. The *scale_pos_weight* parameter, which addresses class imbalance in the dataset, was set to 3.4, calculated as the square root of the ratio of the number of successful quitters to the number of unsuccessful quitters at the time of the wave 6 survey. The model was trained for a specified number of boosting rounds (*n_rounds* = 1000), with early stopping (*early_stopping_rounds* = 400) implemented to cease training if performance did not improve after a certain number of iterations, thus preventing overfitting and conserving computational resources.

Throughout this process, Area Under the Receiver Operating Characteristic Curve [25] (AUC) was used as the evaluation metric to monitor model performance. Averaged AUC and its standard deviation were computed after 1000 runs. Additionally, SHapley Additive exPlanations (SHAP) values [26] were calculated after each iteration to determine the contribution of each variable to the model’s predictions. SHAP, a technique based on Shapley values from cooperative game theory, are used to explain the output of machine learning models [27]. For a given an observation, SHAP values quantify each feature’s contribution to the deviation of the individual prediction of that observation from the average model prediction [27]. Features with positive SHAP values positively influence the prediction, while those with negative values exert a negative impact. By aggregating SHAP values from the 1000 runs, we aimed to gain insights into the directional impact of each instance on the model’s predictions and to identify the top 10 most important variables for predicting participants’ 12-month smoking abstinence status.

### Results

Table 1 presents the basic demographic characteristics of our study sample. The final dataset for analysis includes 6,120 individuals who were current established smokers in wave 5. Of these, 2,329 (38.1%) were aged 18 to 34 years, 2,166 (35.4%) were aged 35 to 54 years, and 1,625 (26.5%) were aged 55 years or older. The sample consists of 3,265 males (53.3%) and 2,854 females (46.7%). In terms of race/ethnicity, 855 (14.0%) identified as Hispanic, 1,002 (16.4%) as non-Hispanic White, 426 (7.0%) as non-Hispanic Black, 3,731 (60.9%) as non-Hispanic other race, and 106 had unspecified ethnicities.

**Table 1:** Basic demographic characteristics of the study sample.

| Characteristics | Wave 5 current established smokers |  |
| --- | --- | --- |
|  | Unweighted number | Unweighted percentage |
| **Overall** | 6120 | 100 |
| **Age** |  |  |
| 18 to 34 years old | 2329 | 38.1 |
| 35 to 54 years old | 2166 | 35.4 |
| 55 or more years old | 1625 | 26.6 |
| **Sex** |  |  |
| Female | 3265 | 53.3 |
| Male | 2854 | 46.7 |
| Missing | 1 | 0.0 |
| **Race/Ethnicity** |  |  |
| Hispanic | 855 | 14.0 |
| Non-Hispanic and Black | 1002 | 16.4 |
| Non-Hispanic and Other | 426 | 7.0 |
| Non-Hispanic and White | 3731 | 61.0 |
| Missing | 106 | 1.6 |
| **Highest education** |  |  |
| >= College | 676 | 11.0 |
| Some college | 2059 | 33.6 |
| ≤ High school/GED | 3353 | 54.8 |
| Missing | 32 | 0.6 |
| **Past 12-month total household income** |  |  |
| >= \$100,000 | 438 | 7.2 |
| \$50,000 - < \$100,000 | 1098 | 17.9 |
| < \$50,000 | 4310 | 70.4 |
| Missing | 274 | 4.5 |

Leveraging only pre-trained knowledge, GPT-4.1 provided robust scores for all 303 variables in predicting 12-month smoking abstinence. Table A2 in the Appendix presents averaged scores and standard deviations of the top 45 variables across 50 different runs. These selected variables encompass various factors, including smoking frequency and habits, social and environmental influences, physical and psychological nicotine dependence, health concerns, cessation attempt and intention, and patterns of other tobacco product usage.

The performance of XGBoost is nearly identical whether considering all 303 wave 5 variables or only the top 45 features selected by GPT-4.1. Specifically, the average AUCs of XGBoost, calculated over 1000 runs, are 0.749 (95% confidence interval: 0.720 - 0.778) when trained on the 303 variables, and 0.752 (95% confidence interval: 0.727 - 0.778) when using the 45 selected features.

Figure 1 presents the top 10 most important predictors of smoking cessation success among 45 selected variables, based on their mean SHAP values - their contribution to the model’s prediction. The SHAP value of each variable for each observation indicates its directional impact on predicting the likelihood of achieving 12-month smoking abstinence. When training XGBoost, 12-month smoking abstinence status is encoded as 1 if an individual reported having not smoked in the past 12 months at time of wave 6 survey, and 0 otherwise. Therefore, variables’ values with negative SHAP values are associated with a decreased likelihood of having not smoked in the past 12 months, while those with positive SHAP values indicate an increased likelihood. For example, Figure 1 shows that individuals with a high “past 30-day cigarette smoking frequency” in wave 5, which indicates greater nicotine dependence, were less likely to achieve 12-month smoking abstinence in wave 6. Individuals who currently used electronic nicotine products every day in wave 5 were highly associated with successfully quitting smoking in wave 6.

**Figure 1:**
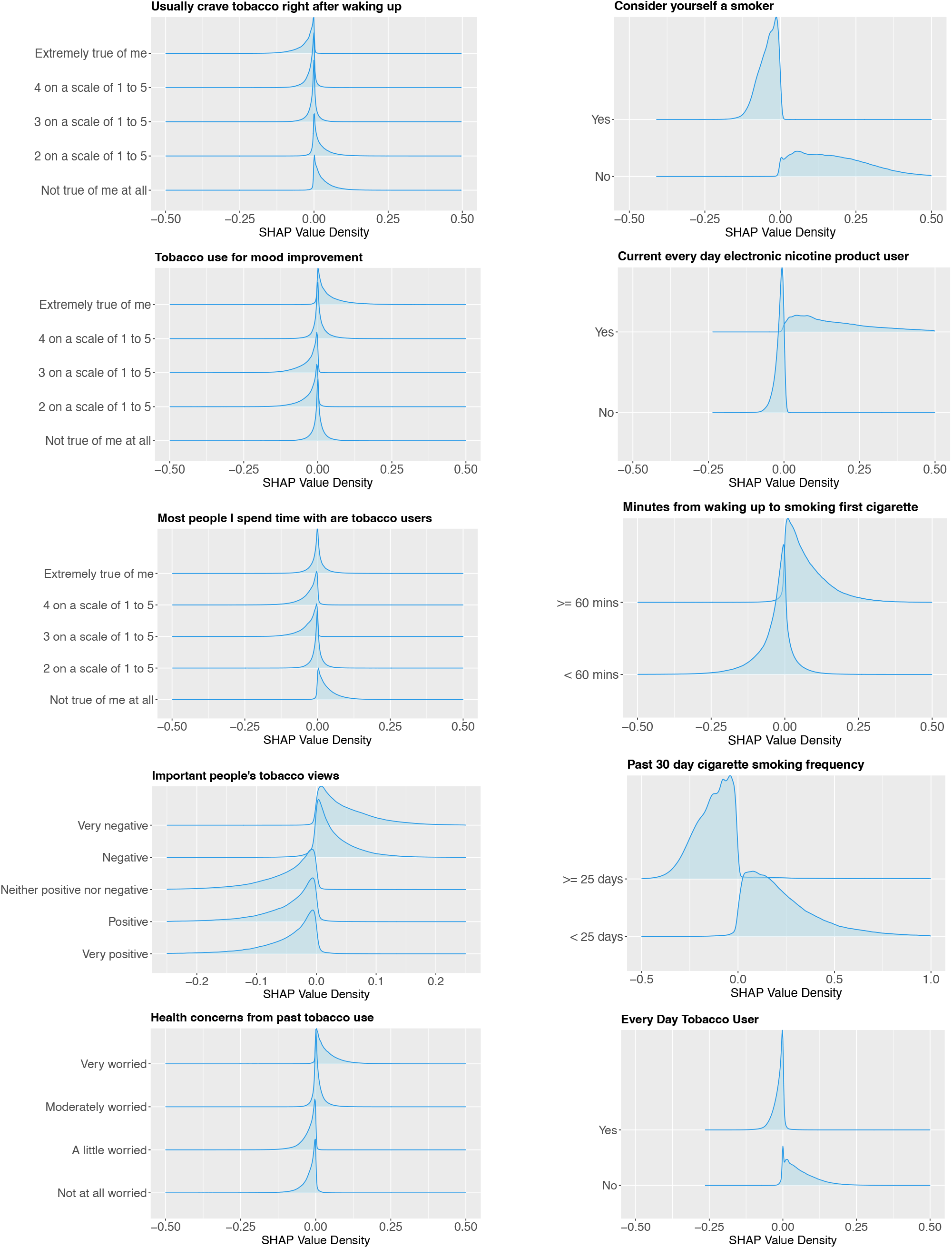
The distribution of SHAP values for the top 10 most important predictors of 12-month smoking abstinence.

## Discussion

To our knowledge, this study is the first to perform text-based feature selection in tobacco control, using GPT-4.1. Leveraging its extensive pre-trained knowledge, GPT-4.1 was able to assign importance scores to wave 5 PATH survey variables for predicting 12-month smoking abstinence status in wave 6, using only variables’ text descriptions and without direct access to the data. With the 45 highest-scored variables, XGBoost predicted smoking cessation status in the outcome wave with high accuracy. In particular, the performance of XGBoost trained with the 45 GPT-4.1-selected variables was similar to that trained with the 303 variables (AUC: 0.752 vs AUC: 0.749). This suggests the high quality of the selected variables, demonstrating the promising capability of GPT-4.1 and LLMs in general in performing text-based feature selection. Based on its provided importance scores, GPT-4.1 can help identify key factors that should be the focus of data collection to address specific research questions. This enables researchers to prioritize this information when designing survey questionnaires.

The list of the top 45 wave 5 variables that are significantly associated with 12-month smoking abstinence in wave 6 encompasses various factors. These include information about participants’ smoking frequency and habits [12, 28, 29], social and environmental influences [8-12, 29], nicotine dependence [30, 31], health concerns [29], cessation attempt and intention [10, 12, 32], and other tobacco product usage [10, 12] (see Table A1). It is worth noting that most of variables used to compute the tobacco dependence score in Strong et. al. [30, 31] are included in our top 45 variables. The association of these diverse factors with successful smoking cessation aligns well with previous research findings. These alignments reinforce the relevance of the selected variables in predicting smoking cessation success, underscoring the efficacy of GPT-4.1’s text-based feature selection.

The SHAP value of each variable for each observation offers valuable information into how individual variables impact the likelihood of achieving 12-month smoking abstinence. The following insights regarding the influence of the top 10 variables are extracted from Figure 1.

Figure 1 indicates that individuals who smoked fewer days in the past 30 days, waited longer to smoke their first cigarette after waking, or usually did not want to smoke/use tobacco products right after waking up were more likely to quit successfully compared to those who smoked more frequently and sooner [12, 28]. In addition, individuals who identified themselves as smokers or current everyday tobacco users (i.e., those who currently used any form of tobacco product, including electronic nicotine products, combustible tobacco products such as traditional cigarettes, and smokeless tobacco products every day) were associated with a lower probability of having abstained from smoking in the past year at the follow-up [12, 32]. It is worth noting that our baseline population of interest consists of individuals who had smoked at least 100 cigarettes in their lifetime and currently smoked cigarettes either some days or every day. This implies that current everyday tobacco users include a subset of our baseline population who smoked every day. These factors, directly or indirectly, reflect individuals’ nicotine dependence levels, indicating that those with lower dependence were more likely to successfully quit smoking within two years [12, 28, 30].

Furthermore, Figure 1 shows that current every day electronic nicotine product users were linked to an increased probability of being 12-month smoking abstinent. The association between daily electronic nicotine product user and smoking cessation success has been controversial in the literature with previous studies reported different findings, see [12, 33, 34] and references therein. More research into this issue is needed to reach a consensus on the role of electronic nicotine products in smoking cessation. Individuals whose important people had negative views on tobacco use and who did not spend time mostly with smokers were more likely to quit smoking compared to their counterparts [10, 35, 36]. This suggests that social environment plays a significant role in smoking cessation. In addition, smokers are moderately or very concerned about their future health due to their tobacco use were linked to an increased likelihood of achieving 12-month smoking abstinence. However, this association has not been well-reported in the literature (for example [29] reported similar finding to ours, while [37] did not). Finally, our findings also indicate that current smokers who completely agreed with the statement “smoking/using tobacco product(s) really helps me feel better if feeling down” in wave 5 were correlated with successfully quitting smoking by wave 6. In summary, further studies are required to verify the associations that are not yet well-established in the literature.

In addition to GPT-4.1, we tested older versions of OpenAI’s GPTs (e.g., GPT-3.5-TURBO and GPT-4-0613). While these models generally performed reasonably well, GPT-4.1 provided more robust importance scores compared to the others. Future studies focusing on testing and comparing different available LLMs would be beneficial in selecting the best model for text-based feature selection. With classical machine learning methods such as Random Forest and XGBoost, we have a clear understanding of the criteria (e.g., SHAP values [13, 14], mean decrease in accuracy and mean decrease impurity [13, 38]) that are used to provide importance scores for variables. However, it is important to note that for LLMs, we do not know the criteria on which their provided scores are based [15]. Because the findings of this study are based on the PATH data, it also bears the limitations of the PATH study [13].

In conclusion, this study highlights the promising capability of LLMs, particularly GPT-4.1, in identifying the most important variables for predicting 12-month smoking abstinence using solely variable descriptions, without direct access to the dataset itself. Leveraging this ability among many others, LLMs can be used to aid researchers in designing survey questionnaires by allowing them to focus on the most relevant factors, thereby enhancing the efficiency and effectiveness of data collection.

## Data Availability

The dataset used in this work is publicly available online.

https://www.icpsr.umich.edu/web/NAHDAP/studies/36498

## Contributors

TTTL, ZZ, KZ, and YH conceptualized the project. TTTL conducted the analysis and wrote the first draft. JY contributed to writing the first draft. WL played a significant role in revising the draft. All the authors contributed to the editing and review of the manuscript.

## Disclaimer

The content is solely the responsibility of the authors and does not necessarily represent the official views of the NIH or the Food and Drug Administration.

## Patient consent for publication

Not required.

## Provenance and peer review

Not commissioned; externally peer reviewed.

