## Supplemental Material for "Identifying Key Predictors of Smoking Cessation Success: Text-Based Feature Selection Using a Large Language Model"

**Prompt template:**

Below is the prompt that we used in this study

Prompt = { "*As a researcher specializing in tobacco cessation behavior analysis, after thoroughly reviewing the literature, your task is to score the importance of the variable,* ***variable_desciption****, in predicting whether an individual will abstain from smoking during the 12-month period preceding their interview, which is scheduled to take place two years from now. Provide a numeric score between 1 and 100 to reflect the variable's importance, with 1 being the least important and 100 being the most important. Ensure your response is formatted precisely as: 'Score: XX', where XX represents your numeric score. Following the score, include a concise reasoning of your rating starting with 'Reasoning:', and limited to one or two sentences.*"}

where ***variable_desciption*** is the description of each variable from Table A1.

**Table A1:** The parameter space of the optimized XGBoost hyperparameters.

| **Hyperparameters** | eta | max_depth | min_child_weight | subsample | colsample_bytree |
|---|---|---|---|---|---|
| **Parameter space** | [0.01, 0.1] | [4L, 25L] | [0, 25] | [0.5, 1] | [0.5, 1] |

**Table A2:** The exhaustive list of the top 45 variables together with their descriptions and their means and standard deviations of important scores provided by GPT-4.1.

| **Category** | **Variable name** | **Variable description** | **Mean score (SD)** | **Mean SHAP value** |
|---|---|---|---|---|
| Smoking Frequency and Habits | R05R_A_NUMDAYS_CIGS | Adult Past 30 Day Cigarette Smoking Frequency | 90 (3) | 0.16220 |
| Nicotine Dependence | R05_AC9022 | Consider yourself a smoker | 85 (2) | 0.05796 |
| Smoking Frequency and Habits | R05R_A_MINFIRST_CIGS | Adult Number of Minutes from Waking Up to Smoking First Cigarette | 85 (1) | 0.05690 |
| Social and Environmental Influences | R05_AX0071 | People who are important to you: Their views on using tobacco | 75 (3) | 0.04514 |
| Social and Environmental Influences | R05_AN0255 | Level of agreement: Most people I spend time with are tobacco users (current established, recent former established or current experimental non-electronic tobacco users) | 81 (2) | 0.02817 |
| Nicotine Dependence | R05_AN0070 | Level of agreement: Smoking/using tobacco product(s) really helps me feel better if feeling down | 78 (1) | 0.02796 |
| Other Tobacco Product Usage Patterns | R05R_A_EDY_EPRODS | Adult Current Every Day Electronic Nicotine Product User | 79 (3) | 0.02557 |
| Health Harm Perceptions and Concerns | R05_AX0105 | Extent to which you are worried that using/your past use of tobacco products will damage your health in the future | 77 (2) | 0.02498 |
| Nicotine Dependence | R05_AN0060 | Level of agreement: Usually want to smoke/use tobacco product(s) right after waking up | 90 (2) | 0.02472 |
| Other Tobacco Product Usage Patterns | R05R_A_EDY_TOB | Adult Every Day Tobacco User | 95 (1) | 0.02469 |
| Enjoying Sensation | R05_AC9045 | Level of Agreement: Enjoy sensation in throat and chest when smoking | 75 (3) | 0.02439 |
| Cessation Efforts and Intentions | R05_AN0105 | In the past 12 months have you tried to quit smoking/using tobacco product(s) | 76 (4) | 0.02389 |

| | | | | |
|---|---|---|---|---|
| Social and Environmental Influences | R05_AR1045 | Statement that best describes rules about smoking a combustible tobacco product inside your home | 76 (3) | 0.02009 |
| Nicotine Dependence | R05_AN0030 | Level of agreement: Urges keep getting stronger if don't/Still have urges to smoke/use tobacco product(s) | 85 (1) | 0.01889 |
| Cessation Efforts and Intentions | R05_AN0235 | Plans to quit smoking/using tobacco product(s) for good | 76 (3) | 0.01829 |
| Social and Environmental Influences | R05_AX0741_10 | People who are important to you use the following products: None of the above | 78 (3) | 0.01730 |
| Nicotine Dependence | R05_AN0100 | Level of agreement: After not smoking/using tobacco product(s) for a while, need to smoke/use tobacco product(s) in order to keep self from experiencing any discomfort | 87 (1) | 0.01693 |
| Smoking Frequency and Habits | R05_AC9053 | Smoke cigarettes more frequently during the first hours after waking compared to rest of the day | 85 (1) | 0.01559 |
| Nicotine Dependence | R05_AN0055 | Level of agreement: Finds self reaching for tobacco product(s) without thinking about it | 79 (2) | 0.01509 |
| Nicotine Dependence | R05_AN0045 | Level of agreement: My tobacco product(s) smoking/use is out of control/My urge to smoke/use tobacco product(s) is out of control | 83 (2) | 0.01479 |
| Nicotine Dependence | R05_AN0065 | Level of agreement: Can only go a couple of hours without smoking/using tobacco product(s) | 86 (1) | 0.01478 |
| Nicotine Dependence | R05_AN0035 | Level of agreement: Tobacco product(s) control me | 79 (2) | 0.01457 |
| Nicotine Dependence | R05_AN0010 | Consider yourself to be addicted to tobacco product(s) | 76 (3) | 0.01429 |
| Nicotine Dependence | R05_AN0095 | Level of agreement: After not smoking/using tobacco product(s) for a while, need to smoke/use tobacco product(s) in order to feel less restless and irritable | 86 (2) | 0.01320 |
| Nicotine Dependence | R05_AN0085 | Level of agreement: Would find it really hard to stop smoking/using tobacco product(s) | 84 (1) | 0.01258 |
| Nicotine Dependence | R05_AN0050 | Level of agreement: Frequently smoke/use tobacco product(s) without thinking about it | 83 (2) | 0.01168 |
| Other Tobacco Product Usage Patterns | R05R_A_SDY_TOB | Adult Some Day Tobacco User | 81 (3) | 0.01110 |
| Nicotine Dependence | R05_AN0025 | Level of agreement: Frequently crave tobacco product(s) | 85 (1) | 0.01098 |
| Nicotine Dependence | R05_AN0015 | Has strong cravings to smoke/use tobacco product(s) | 85 (1) | 0.01084 |
| Health Harm Perceptions and Concerns | R05_AX0104 | Extent to which using/past use of tobacco products damaged your health | 77 (3) | 0.01044 |
| Nicotine Dependence | R05_AN0090 | Level of agreement: Would find it hard to stop smoking/using tobacco product(s) for a week | 76 (3) | 0.01014 |
| Nicotine Dependence | R05_AC9054 | Smoke cigarettes even if ill in bed all day | 85 (2) | 0.00839 |

| | | | | |
|---|---|---|---|---|
| Health Harm Perceptions and Concerns | R05_AN0110 | Do you believe that smoking/using tobacco products is causing/caused a health problem or made it worse | 82 (3) | 0.00723 |
| Nicotine Dependence | R05_AN0020 | Felt like you really needed to smoke/use tobacco product(s) | 85 (1) | 0.00634 |
| Other Tobacco Product Usage Patterns | R05R_A_EDY_CIGAR | Adult Current Every Day Cigar Smoker | 85 (2) | 0.00575 |
| Other Tobacco Product Usage Patterns | R05R_A_P30D_GRILLO | Adult Past 30 Day Cigarillo Smoker | 77 (4) | 0.00481 |
| Other Tobacco Product Usage Patterns | R05R_A_CUR_EXPR_TOB | Adult Current Experimental Tobacco User | 85 (1) | 0.00355 |
| Other Tobacco Product Usage Patterns | R05R_A_P12M_GTRAD | Adult Past 12 Month Traditional Cigar Smokers | 79 (4) | 0.00350 |
| Other Tobacco Product Usage Patterns | R05R_A_CUR_EXPR_EPRODS | Adult Current Experimental Electronic Nicotine Product User | 78 (2) | 0.00157 |
| Other Tobacco Product Usage Patterns | R05R_A_P30D_GTRAD | Adult Past 30 Day Traditional Cigar Smoker | 77 (4) | 0.00130 |
| Other Tobacco Product Usage Patterns | R05R_A_CUR_EDSD_GFILTR | Adult Current Every Day/Some Day (Without Threshold) Filtered Cigar Smoker | 75 (4) | 0.00127 |
| Other Tobacco Product Usage Patterns | R05R_A_SDY_EPRODS | Adult Current Some Day Electronic Nicotine Product User | 77 (3) | 0.00124 |
| Other Tobacco Product Usage Patterns | R05R_A_CUR_EDSD_HOOK | Adult Current Every Day/Some Day (Without Threshold) Hookah Smoker | 76 (3) | 0.00110 |
| Other Tobacco Product Usage Patterns | R05R_A_CUR_EDSD_GTRAD | Adult Current Every Day/Some Day (Without Threshold) Traditional Cigar Smoker | 77 (3) | 0.00080 |
| Other Tobacco Product Usage Patterns | R05R_A_CUR_ESTD_GTRAD | Adult Current Established Traditional Cigar Smoker | 78 (4) | 0.00009 |
